# Development and Validation of a Machine Learning Model to Predict Prognosis in Patients with Advanced Head and Neck Cancer

**DOI:** 10.64898/2026.05.27.26354194

**Authors:** Kevin Zhang, Lan Gao, Daniel John, Wei Tse Li, Michael Hogarth, Charles S. Coffey, Weg M. Ongkeko

## Abstract

**Importance:** Prognostic tools beyond staging are needed to guide treatment and counseling in head and neck squamous cell carcinoma (HNSCC).

**Objective:** To develop and externally validate a machine learning model predicting survival in advanced HNSCC using routinely collected clinical and biomarker data.

**Design, Setting, and Participants:** Retrospective, multi-institutional cohort study including 2,385 patients with stage III–IV HNSCC diagnosed from 2012–2022 in the University of California Health Data Warehouse (UCHDW). Patients were randomly split into training (n = 1,908) and test (n = 477) sets. Partial external validation used 7,749 patients from the Surveillance, Epidemiology, and End Results (SEER) registry (2010–2020).

**Exposures:** Demographic, tumor, treatment, comorbidity, and biomarker variables recorded at or before diagnosis.

**Main Outcomes and Measures:** The primary outcome was all-cause mortality within 70 months. Cox proportional hazards models included all predictors. Discrimination was assessed with Harrell’s concordance index (C-index), calibration with predicted vs observed survival, and stratification with Kaplan–Meier curves. A Random Survival Forest (RSF) was trained for benchmarking and interpretability using Shapley Additive exPlanations (SHAP).

**Results:** Among 2,385 patients in UCHDW (median age, 63 years; 29.0% mortality), the Cox model achieved a C-index of 0.735 in the internal test set. Risk quartiles showed clear separation on Kaplan–Meier curves (log-rank p < 0.0001). In the SEER cohort (n = 7,749), where only demographic, staging, subsite, and treatment variables were available, the reduced Cox model achieved a C-index of 0.688, with calibration showing modest underestimation of survival in high-risk groups. Age, T stage, Charlson Comorbidity Index, neutrophil-to-lymphocyte ratio, and platelet count were among the strongest predictors, while surgery was associated with improved survival. The RSF achieved a C-index of 0.758 internally, with SHAP highlighting nonlinear effects of albumin, BMI, and inflammatory markers.

**Conclusions and Relevance:** A machine learning model using routine clinical and biomarker data demonstrated good prognostic performance in advanced HNSCC, with partial external validation. Such approaches may support individualized survival estimates, risk stratification, and treatment discussions, but broader validation is required before clinical adoption.

**Key Points:** *Question:* Can a machine learning model accurately predict prognosis in patients with head and neck cancer utilizing only clinical features and biomarkers?

*Finding:* In a study of 2,385 patients with advanced HNSCC from the UC Health Data Warehouse, a Cox model incorporating demographics, staging, comorbidities, treatment, and clinical biomarkers showed good internal discrimination (C-index 0.735). In partial external validation using 7,749 SEER patients with only demographic, staging, subsite, and treatment variables, performance remained acceptable (C-index 0.688) with preserved risk stratification.

*Meaning:* Machine learning using clinical data can predict prognosis in head and neck cancer; this tool could be used to provide individualized survival estimates and help guide treatment discussions and risk stratification at diagnosis.

## Introduction

Head and neck squamous cell carcinoma (HNSCC) is the seventh most common cancer worldwide, with around 890,000 new cases and 450,000 deaths every year [Barsouk et al., 2023; Bray et al., 2018; Ferlay et al., 2019]. Given this global burden, accurately predicting patient outcomes is essential for guiding treatment and counseling.

Traditional HNSCC prognosis assessment includes evaluating histopathologic features and patient TNM stages. The conventional method inadequately captures the diverse clinical behaviors of HNSCC, with patients at the same stage often experiencing different outcomes [Chen et al., 2022]. Moreover, physicians often provide inaccurate survival estimates [Hoesseini et al., 2020]. Evidently, there is a need for a more objective, precise prognostic tool that incorporates more factors to support decision-making in HNSCC.

In this study, we used machine learning to incorporate complex patient data into an accurate prognostic model, as it has been shown to improve the prediction in oncology and other diseases [(Hoesseini et al., 2021), (Li et al., 2020), (Diaz Badilla et al., 2025)]. Unlike conventional TNM staging or physician estimates, ML can integrate diverse and clinically relevant variables, such as demographics, BMI [(Pandita, 2023)], comorbidities [(Hogarth et al., 2023), (Paleri et al., 2010)], biomarkers [(Johnson et al., 2020), (Peng et al., 2024)], and treatment details [(Tian et al., 2025)].

Thus, we sought to develop and validate an ML-based prognostic model that predicts survival probability for patients with HNSCC. By providing more objective and individualized estimates, this tool aims to overcome the limitations of current prognosis assessment and support better clinical decision-making.

## Methods

### Study Design and Data Sources

This retrospective, multi-institutional cohort study followed TRIPOD reporting guidelines (Type 3) [Collins et al., 2015]. Data came from the University of California Health Data Warehouse (UCHDW) and the Surveillance, Epidemiology, and End Results (SEER) program.

The UCHDW is a HIPAA-compliant repository of deidentified clinical data from six UC health systems, harmonized to the Observational Medical Outcomes Partnership (OMOP) common data model [Hripcsak et al., 2015] and curated by the Center for Data-Driven Insights and Innovation. As of 2025, it contains >1.2 million patients with encounters since 2012.

The SEER validation cohort used publicly available, deidentified data from the 2010–2020 registry [Doll et al., 2018], which covers ∼28% of the U.S. population. Eligible patients were adults (≥18 years) with stage III–IV HNSCC, identified by ICD-O-3 site codes and SEER summary stage. Patients identified only by death certificate or autopsy or with missing survival despite being listed as alive were excluded. Data were extracted with SEER*Stat (v8.3.5). Use of this dataset was exempt from institutional review board oversight.

### Study Population

Eligible patients were adults aged 18 years or older with a new diagnosis of stage III or IV head and neck squamous cell carcinoma (HNSCC). Diagnoses were identified using International Classification of Diseases for Oncology, Third Edition (ICD-O-3) site codes and clinical staging from the American Joint Committee on Cancer (AJCC) 7th edition [Zanoni et al., 2019]. Patients were excluded if they had missing survival time or unknown staging information. For patients without a recorded death event, survival time was censored at the date of last known follow-up, defined as the most recent visit date within the EHR. Patients without any post-diagnosis follow-up data were excluded.

The final development cohort from the UCHDW included 2,385 patients, with 1,908 assigned to the training set and 477 to the internal test set. In this cohort, 38.2% of patients experienced death during follow-up. The external validation cohort from SEER included 7,749 patients with advanced-stage HNSCC meeting the same inclusion criteria. Our patient selection process is further detailed in Supplemental Figure 1.

**Supplemental Figure 1.**
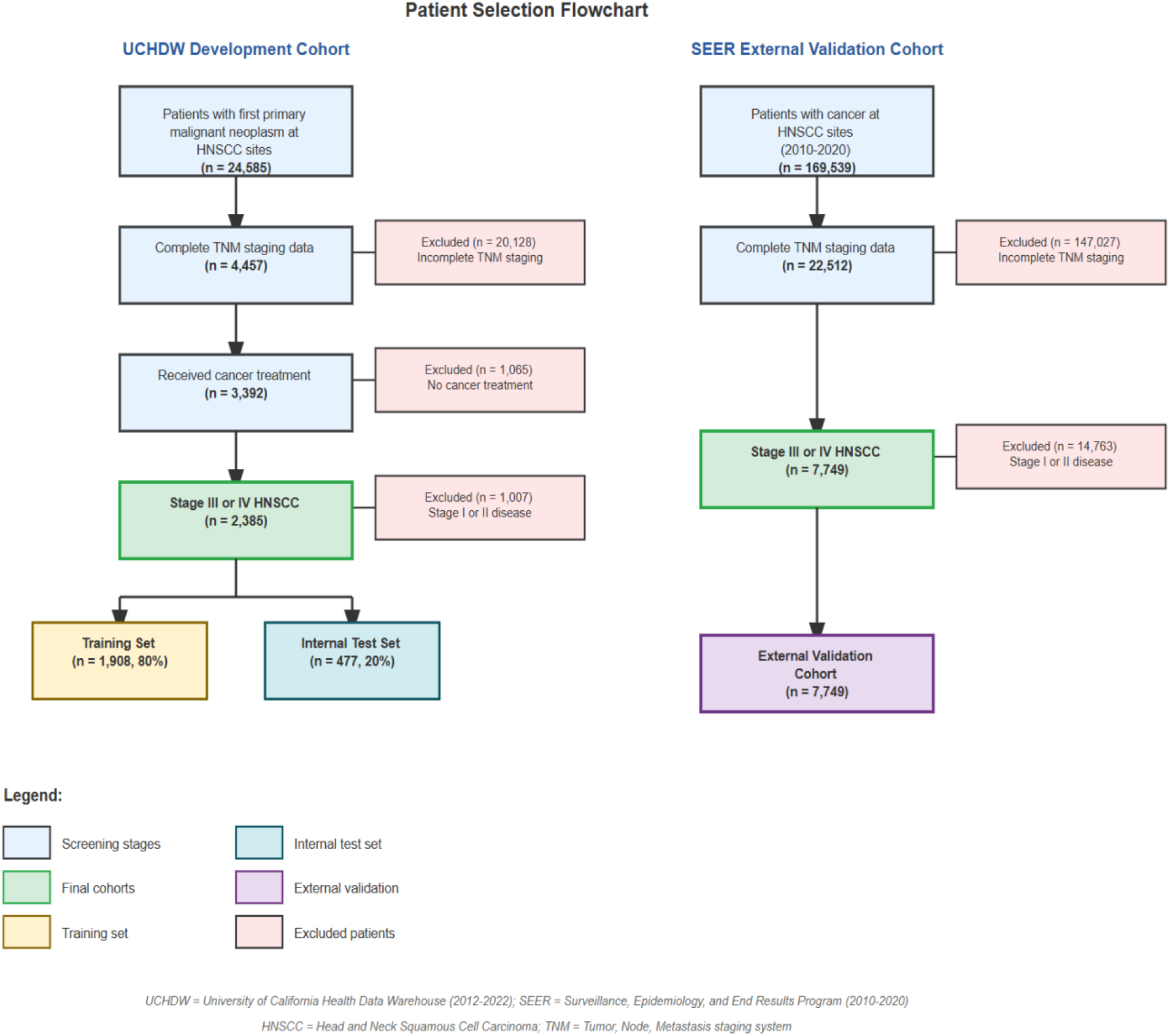
Patient Inclusion Flowchart.

### Outcome Definition

The primary outcome was time to all-cause death, measured in months from diagnosis. Patients were administratively censored at 70 months or at the time of last follow-up, whichever came first. Death status was ascertained through UCHDW for the development cohort and SEER mortality files for the validation cohort. Patients with neither a death event nor post-diagnosis follow-up were excluded from analysis.

### Predictors

All predictors were defined using information available at the time of diagnosis and selected for their clinical relevance and prior evidence of prognostic value in head and neck cancer. Demographic variables included age at diagnosis (continuous) and sex (binary). Tumor characteristics comprised primary tumor subsite (such as tonsil, base of tongue, or larynx) and clinical T, N, and M stage according to the AJCC 7th edition [Edge & Compton, 2010]. Treatment variables captured binary indicators for surgery, radiation therapy, and chemotherapy. Biomarkers included serum albumin [Lim et al., 2017], platelet count [Pardo et al., 2017], body mass index (BMI) [Hodbay et al., 2023], and neutrophil-to-lymphocyte ratio (NLR) [Yang et al., 2019]. Comorbidity burden was quantified using a modified Charlson Comorbidity Index [Ruud et al., 2021] derived from structured fields in the electronic health record (EHR). All demographic, tumor, biomarker, and comorbidity variables were restricted to information recorded prior to or at the time of diagnosis, while treatment details reflected therapies received after diagnosis but before outcome ascertainment. All data were extracted from structured EHR fields within the UC Health Data Warehouse.

#### Missing Data

Missing data were generally infrequent, with the exception of selected biomarkers. Neutrophil-to-lymphocyte ratio (NLR) was missing in 36.8% of patients, and serum albumin was missing in 12.0%. For continuous variables with low levels of missingness, median imputation was used. For variables with higher missingness, such as NLR, we applied k-nearest neighbors (k-NN) imputation and included a missingness indicator variable in the model[Beretta et al., 2016]. Categorical variables with missing values were also assigned a separate missing indicator category. All predictors were retained given their clinical relevance.

### Model Development

A Cox proportional hazards model [Cox et al., 1972] was trained on 80% of the development cohort (n = 1,908). The Cox model estimates the hazard function as:

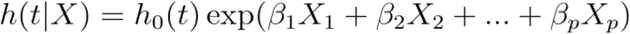

where *h*(*t* | X) is the hazard at time *t* for a patient with covariate vector (*X* = *X*_1_, *X*_2_,…, *X*_*p*_),*h*_0_(*t*) is the baseline hazard function, and *β = β*_1,_ *β*_2_,…, *β*_*p*_) are the regression coefficients estimated from the training data.

The resulting model coefficients were used to compute a prognostic index (PI) for each patient in the internal test and external validation sets:

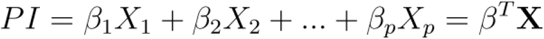

A higher PI indicates greater predicted risk. Hazard ratios (HR) from the Cox model quantify the multiplicative change in hazard associated with a one-unit increase in each predictor; for example, an HR of 1.32 for age indicates that each standard deviation increase in age is associated with a 32% increase in the hazard of death, holding other variables constant. This PI was then used to estimate predicted survival probabilities via the survival function:

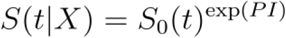

Where *S*_0_(*t*) is the baseline survival function at time *t*. Patients were stratified into quartiles based on their PI values for Kaplan-Meier survival analysis [Kaplan et al., 1958].

Additionally, a random survival forest (RSF) model [Ishwaran et al., 2008] was trained using 500 trees, log-rank splitting, and a minimum node size of 15. Hyperparameters were tuned using grid search with 5-fold cross-validation. The RSF was used for internal benchmarking only and was not externally validated. Its primary purpose was to explore potential nonlinear relationships and interactions and to assess feature importance using SHapley Additive exPlanations (SHAP) [Lundberg et al., 2017] values derived from predicted risk scores.

The internal test set (n = 477) was used to assess model discrimination with Harrell’s concordance index (C-index) [Harrell et al., 1996]. The C-index measures the probability that, for a randomly selected pair of patients, the patient who experienced the event first has a higher predicted risk. Values range from 0.5 (no discrimination, equivalent to random chance) to 1.0 (perfect discrimination), with values >0.7 generally considered acceptable for clinical prediction models. Stratification ability was examined by plotting Kaplan–Meier survival curves across quartiles of predicted risk.

For external validation, the final Cox model was applied to the SEER cohort using only predictors available in both datasets. Laboratory-based biomarkers (e.g., albumin, NLR, platelets) and comorbidity indices were not available in SEER and were excluded. The external validation model therefore included demographic, staging, subsite, and treatment variables. Calibration at 70 months was evaluated in the SEER cohort by comparing predicted risk with Kaplan–Meier estimated survival within deciles of predicted risk.

To support clinical interpretability, we applied SHapley Additive exPlanations (SHAP) [Lundberg & Lee, 2017] to the predicted risk scores generated by the random survival forest (RSF) model. SHAP values provide a patient-level decomposition of model predictions, enabling insight into individual feature contributions. We used TreeExplainer from the SHAP Python package to compute SHAP values on the RSF-derived risk scores. Stratified sampling was applied to ensure equal representation of events and non-events in the interpretability subset. Analyses were conducted in Python 3.11 using the lifelines, scikit-survival, and SHAP libraries.

## Results

The development cohort included 2,385 patients with stage III–IV HNSCC, of whom 1,908 (80%) were assigned to the training set and 477 (20%) to the internal test set. Baseline demographics, tumor characteristics, comorbidities, and treatment distributions are summarized in Table 1. The median ages at diagnosis were 63 (training) and 64 (test). During follow-up, 29.2% and 28.6% of patients died in the training and test sets, respectively.

**Table 1.**
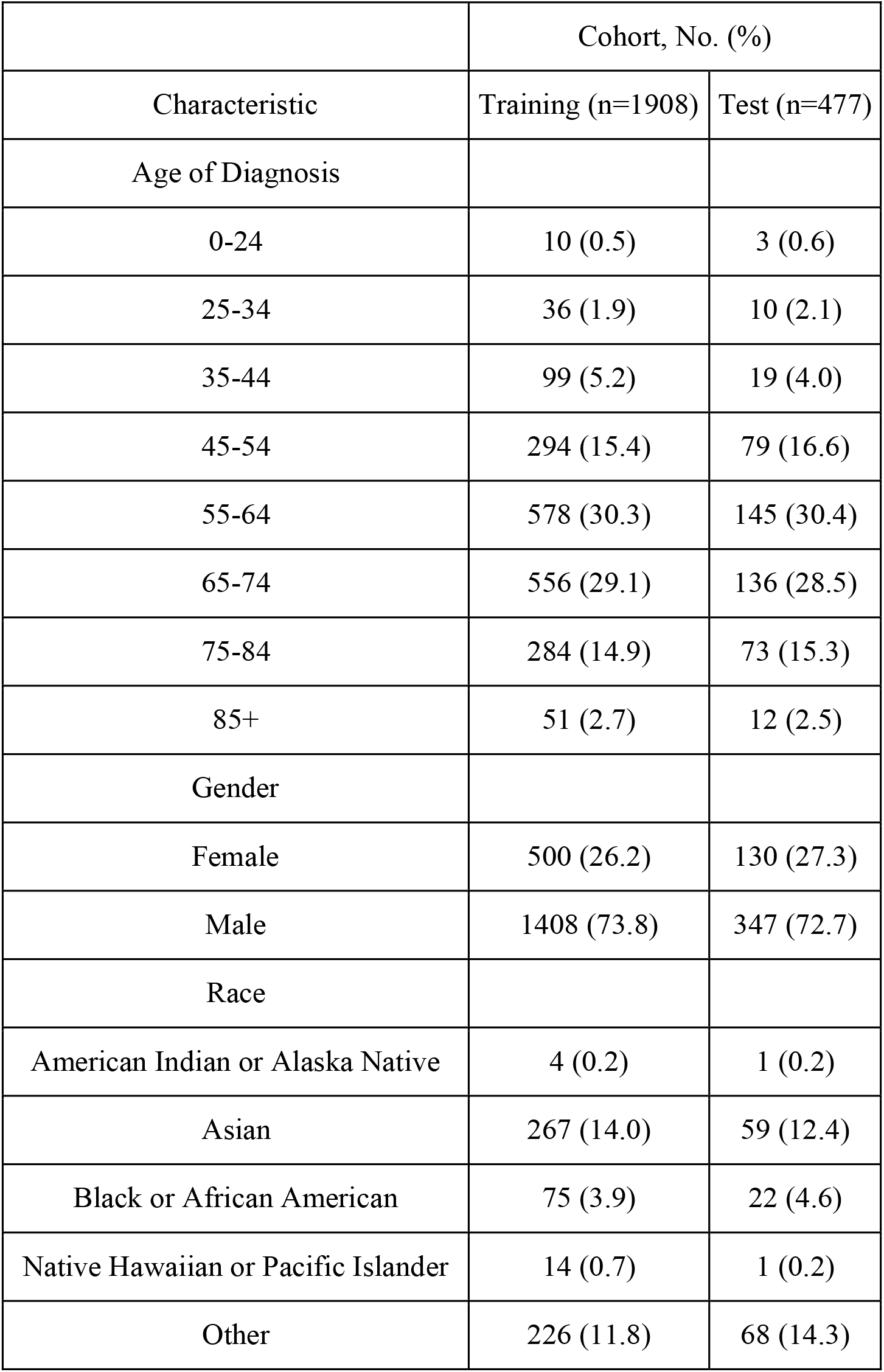
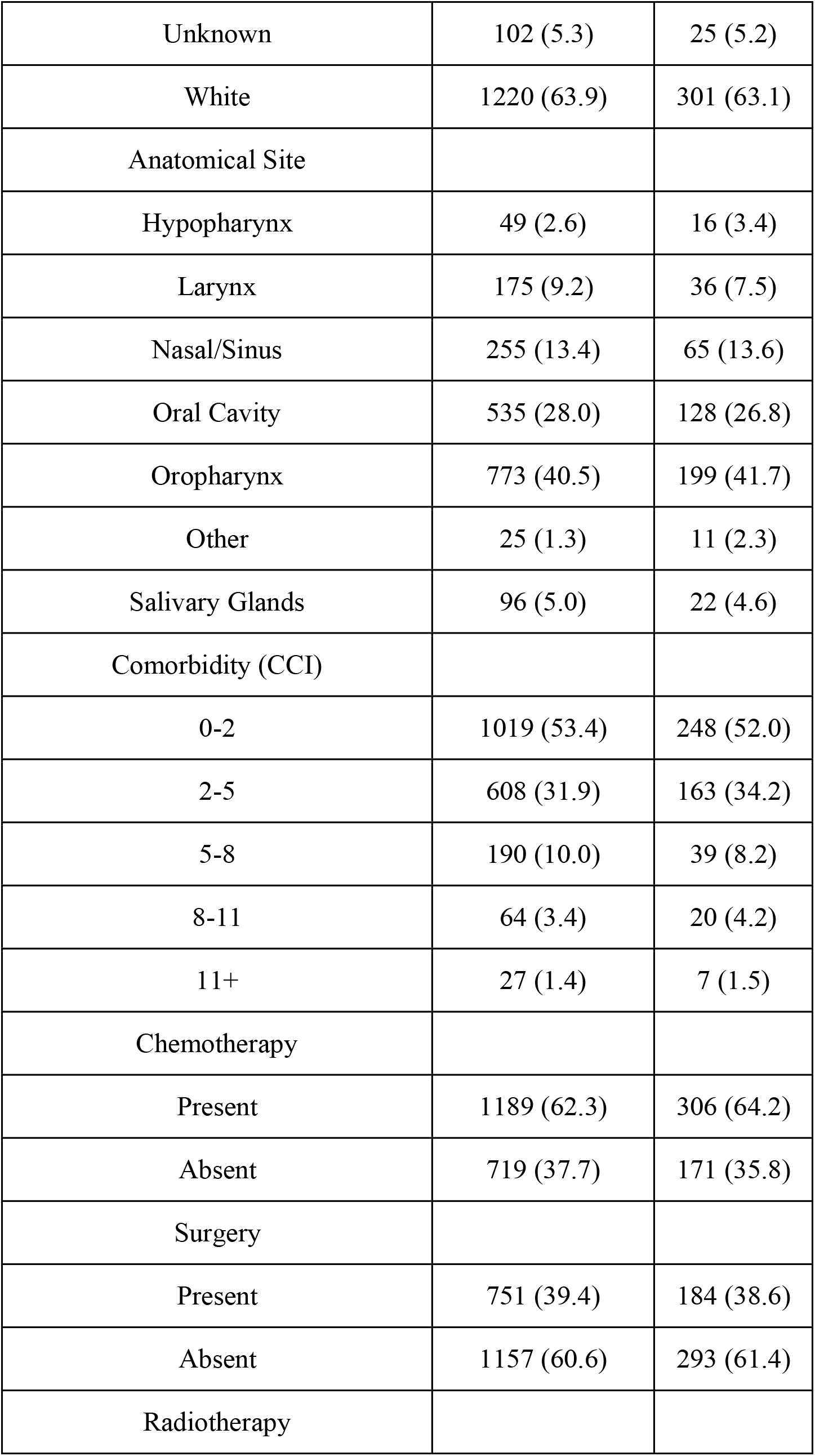
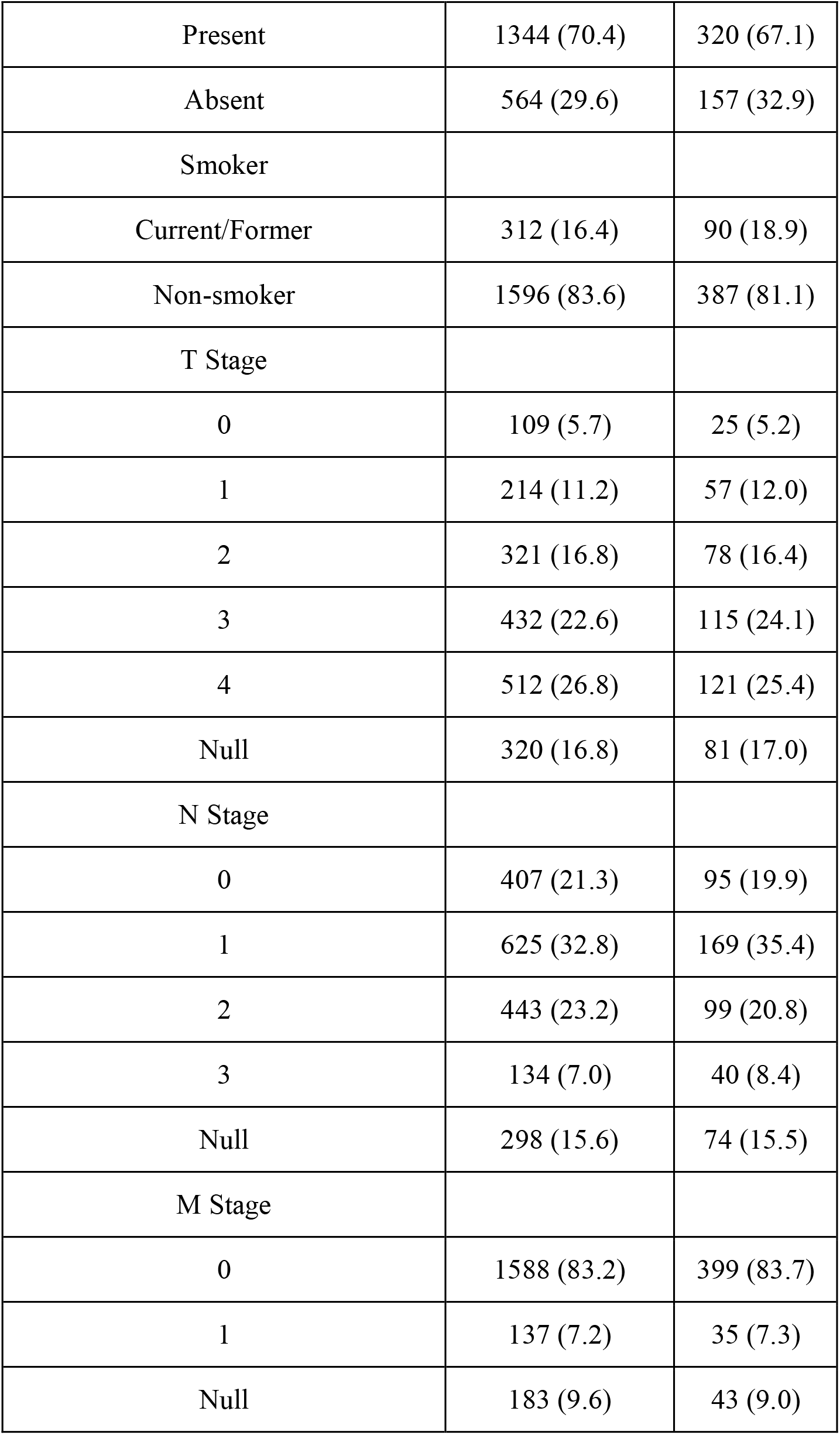
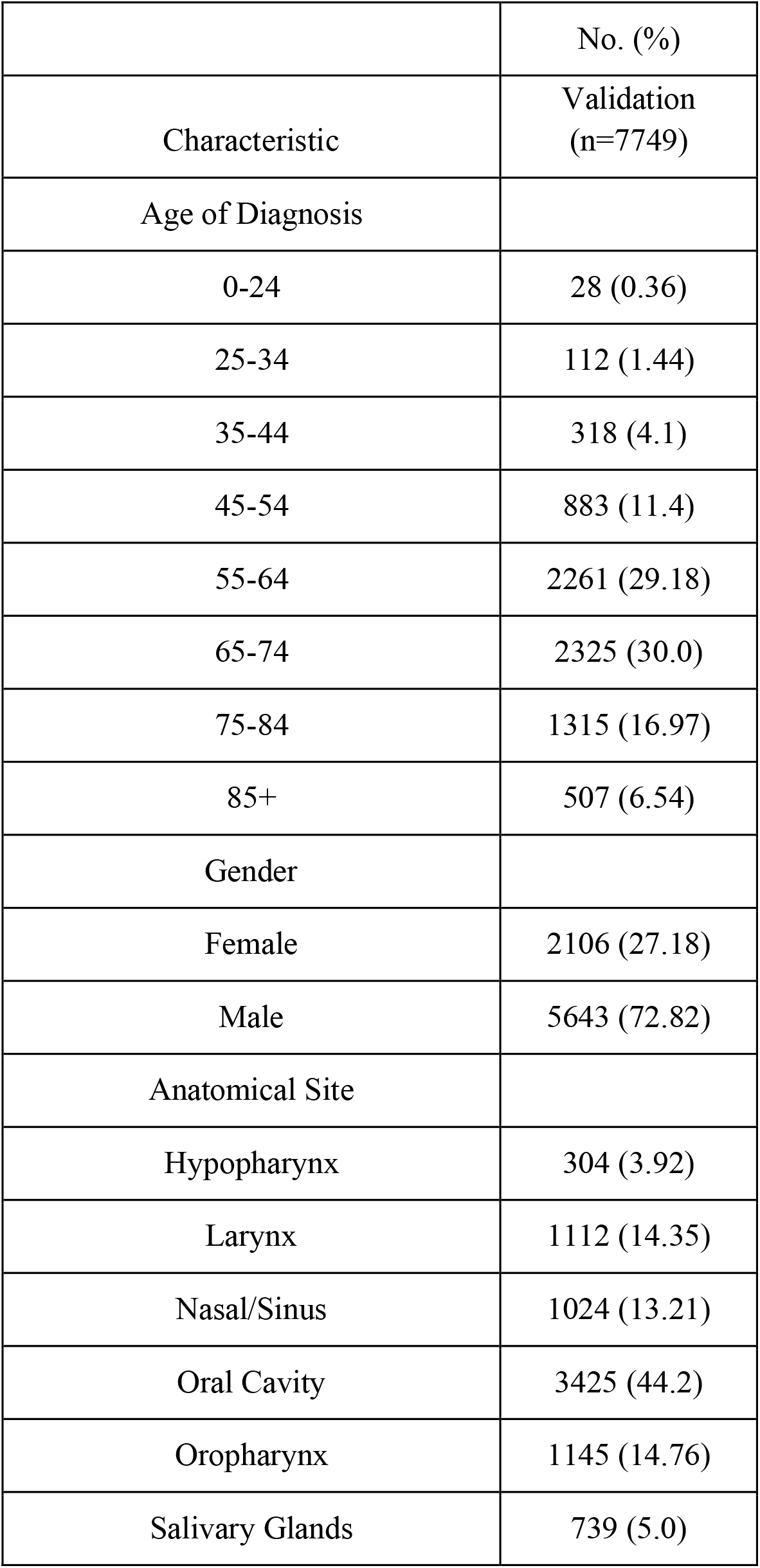
Demographics and Clinical Characteristics by Training and Testing Cohorts.

**Table 1.**
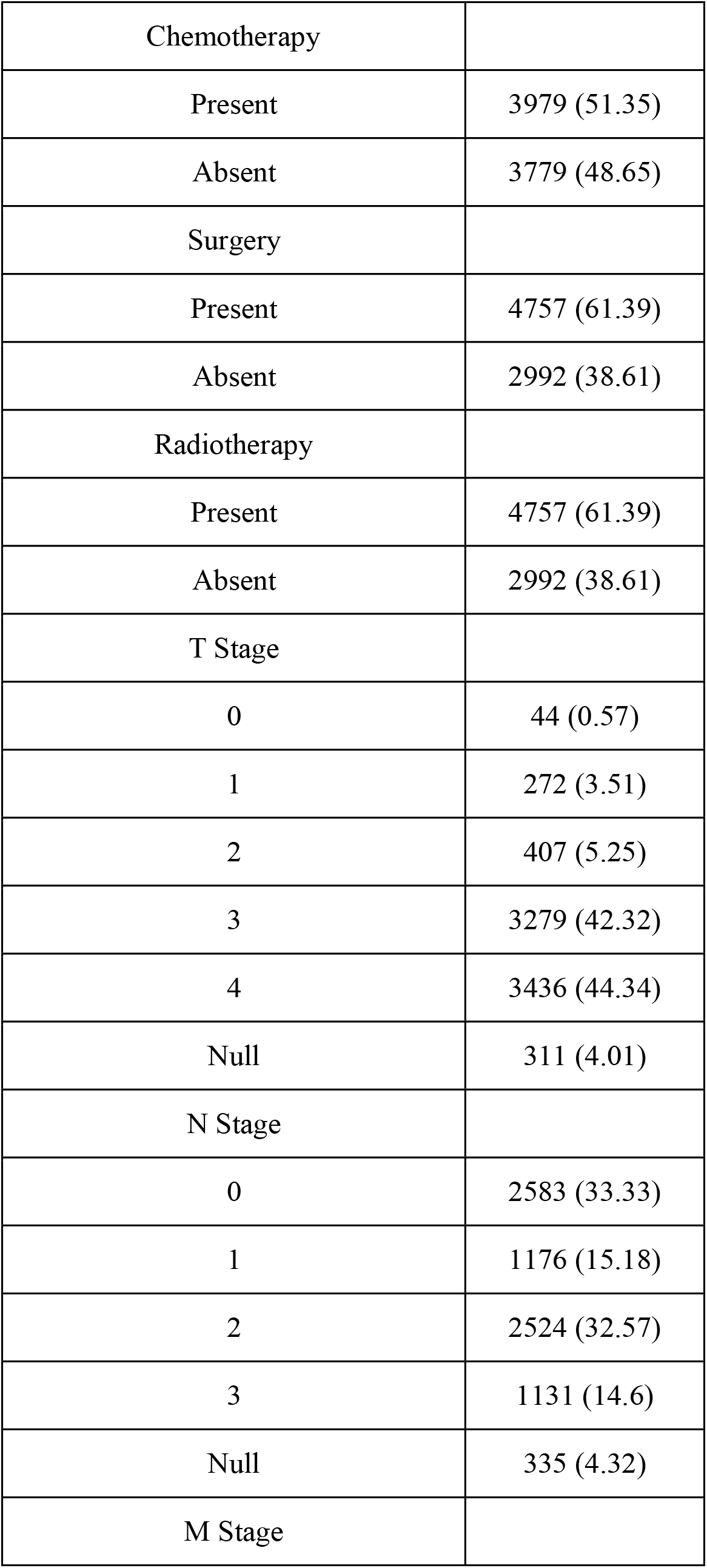
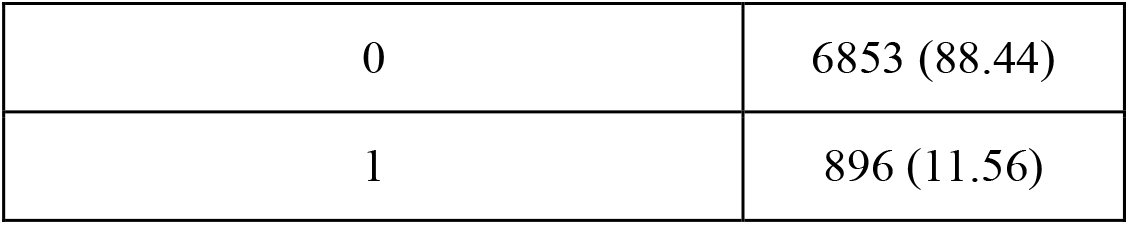
Panel 2. Demographics and Clinical Characteristics, SEER Validation Cohort.

A Cox proportional hazards model was fit using all candidate predictors (Figure 1). Age (HR 1.32, 95% CI 1.20–1.46) and clinical T stage (HR 1.39, 95% CI 1.25–1.54) were most strongly associated with increased mortality. Higher Charlson Comorbidity Index (HR 1.20, 95% CI 1.10–1.30), platelet count (HR 1.18, 95% CI 1.08–1.29), and neutrophil-to-lymphocyte ratio (HR 1.17, 95% CI 1.13–1.22) were also associated with worse outcomes. Surgery was associated with improved survival (HR 0.86, 95% CI 0.78–0.95), while chemotherapy and radiation were not significant after adjustment. Compared with hypopharynx (reference), oropharyngeal tumors had lower risk (HR 0.66, 95% CI 0.53–0.81), with smaller reductions seen for oral cavity, larynx, and nasal/sinus sites.

**Figure 1.**
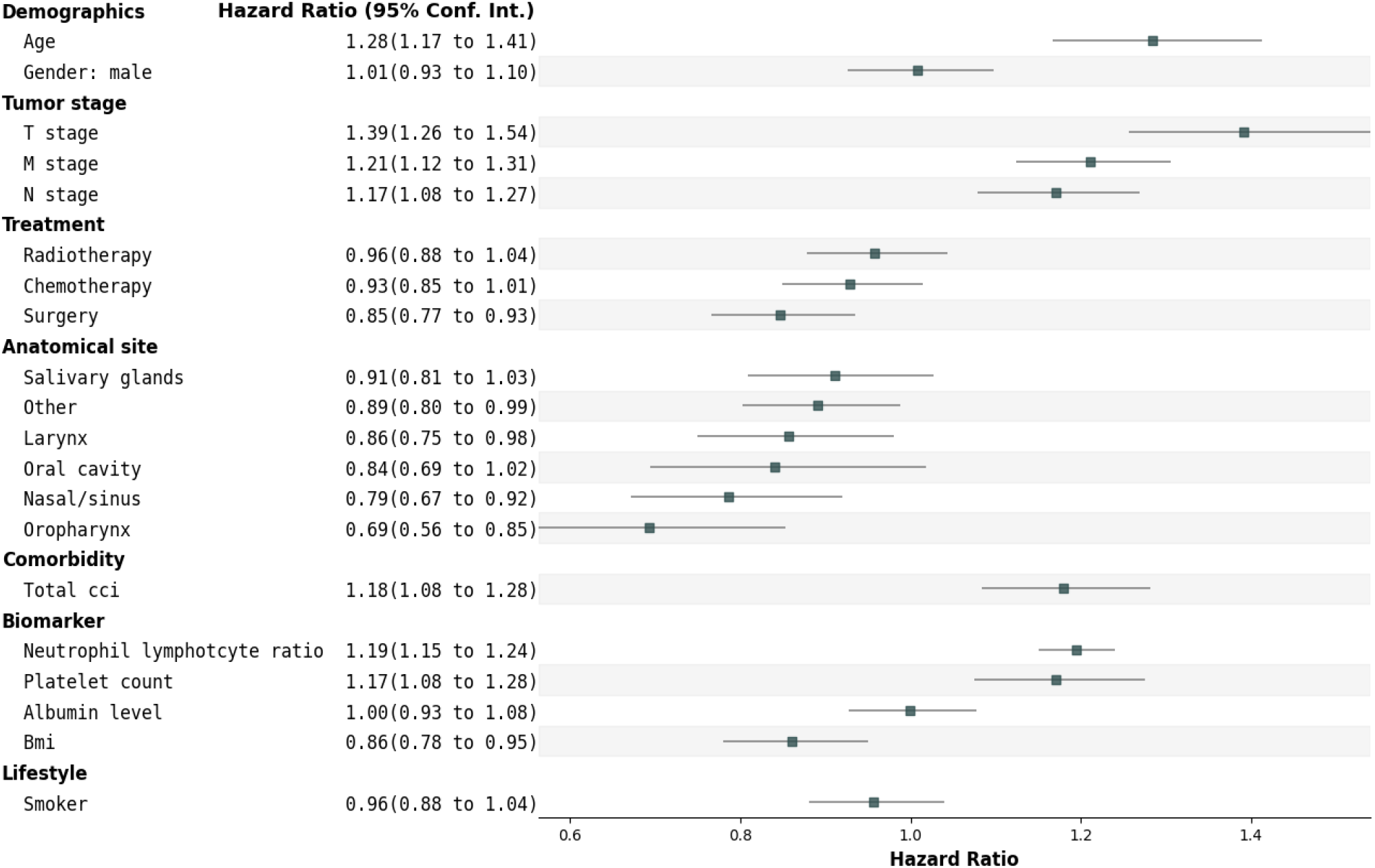
Forest Plot of Covariates for the Cox Proportional Hazards Model. Forest plot of hazard ratios (HRs) with 95% CIs for clinical and biomarker predictors of overall survival in 2,385 patients with stage III–IV head and neck squamous cell carcinoma. Hypopharynx served as the reference category for anatomic site.

To evaluate overall model performance, we calculated Harrell’s C-index and examined survival curves stratified by the prognostic index. On the test set, the Cox model achieved a C-index of 0.735. Kaplan–Meier curves stratified by prognostic index quartiles demonstrated clear separation (log-rank p < .0001) (Figure 2).

**Figure 2.**
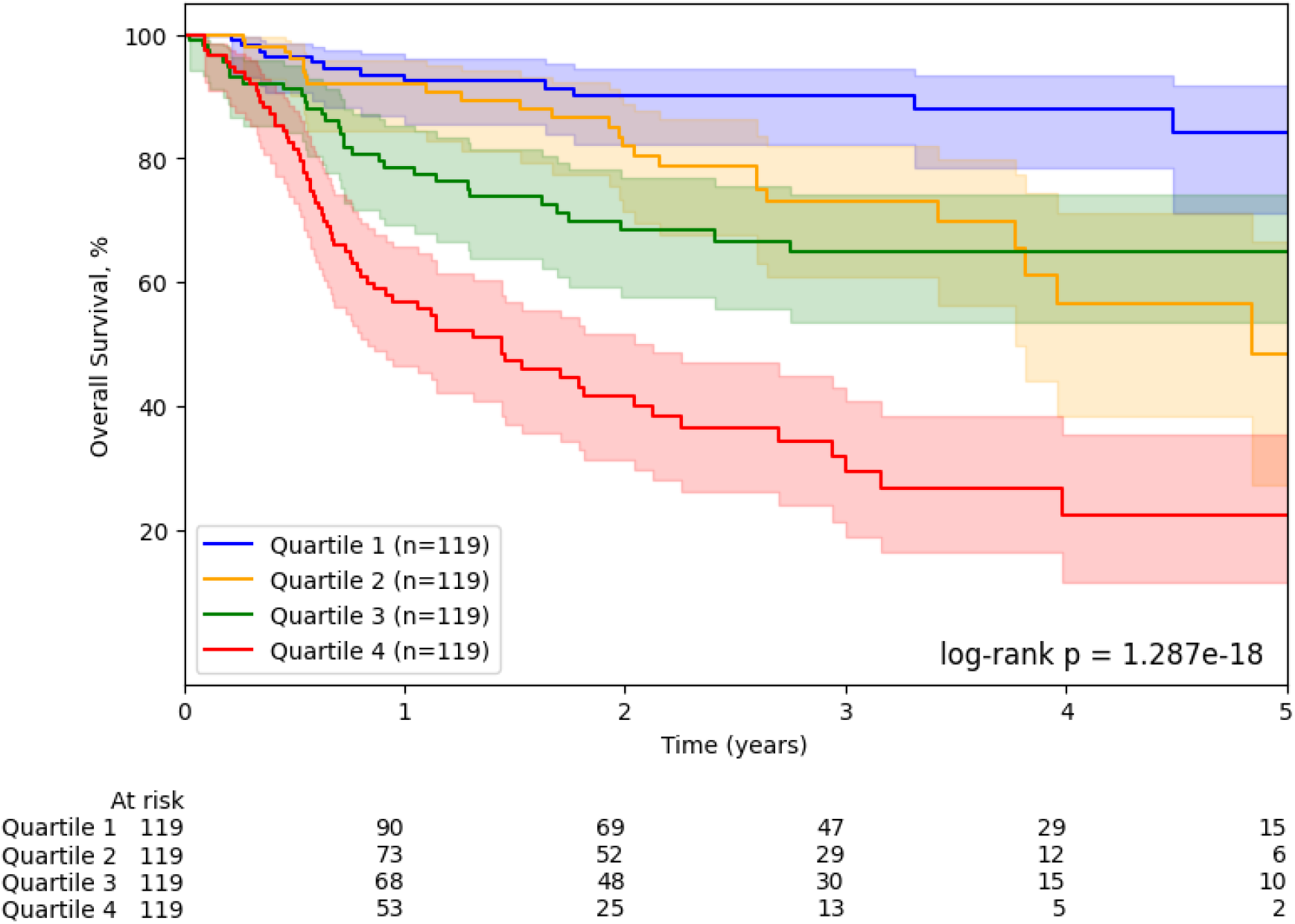
Kaplan–Meier Curves Stratified by Prognostic Index Quartiles. Overall survival in the internal test set (n = 477) according to quartiles of the Cox model prognostic index, with 95% CI bands and p-value.

The RSF model achieved a C-index of 0.758 on the test set. In the RSF, features with the highest average SHAP contributions were age, clinical T stage, Charlson Comorbidity Index, platelet count, and neutrophil-to-lymphocyte ratio. Albumin also ranked among the top contributors, despite a lack of significance in the Cox Model. A SHAP summary plot is shown in Figure 3.

**Figure 3.**
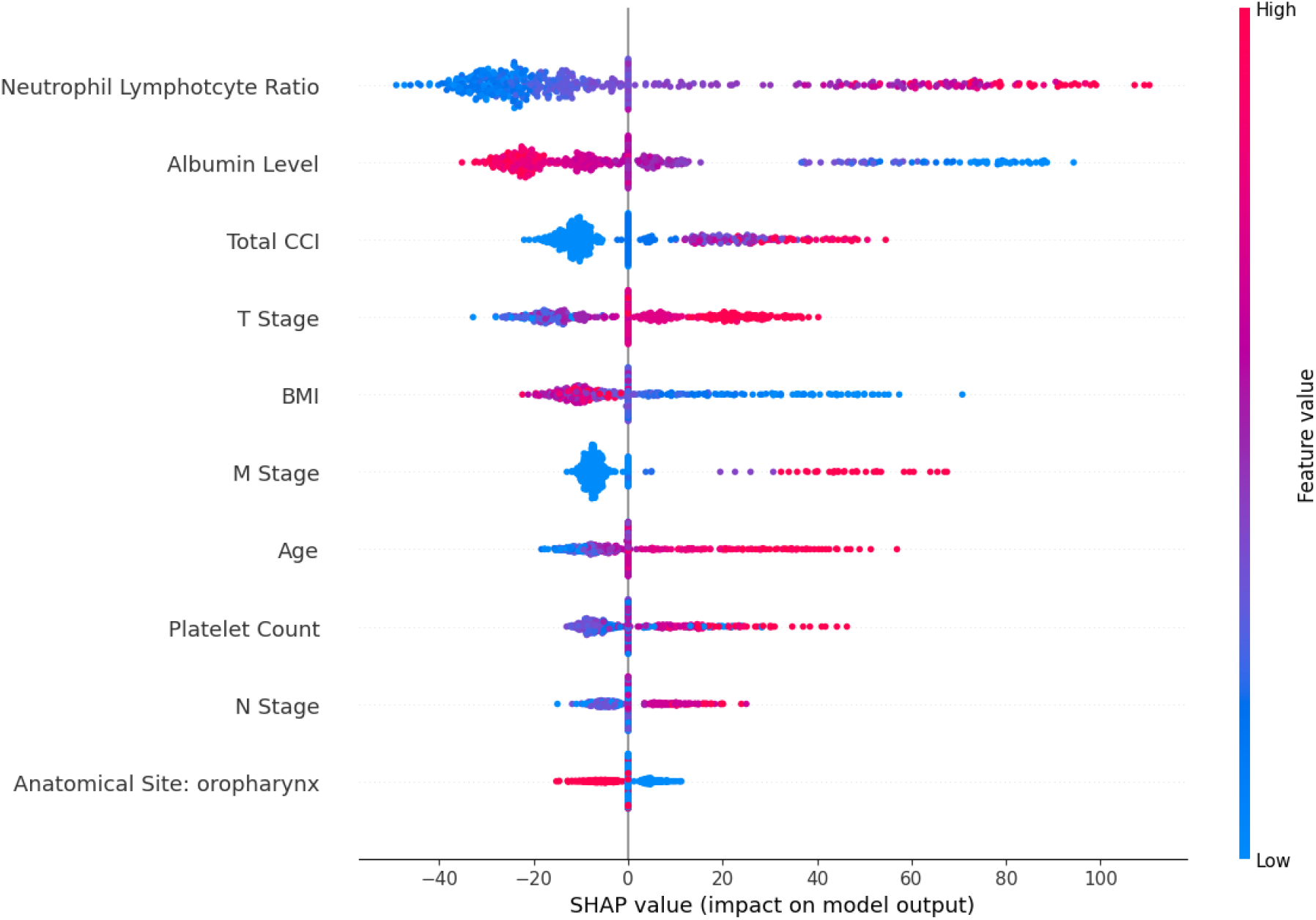
SHAP (SHapley Additive exPlanations) Summary Plot: Feature importance ranked by mean SHAP value in the random survival forest, demonstrating the relative contribution of clinical and biomarker variables to predicted risk. Top 10 features included.

**Figure 4.**
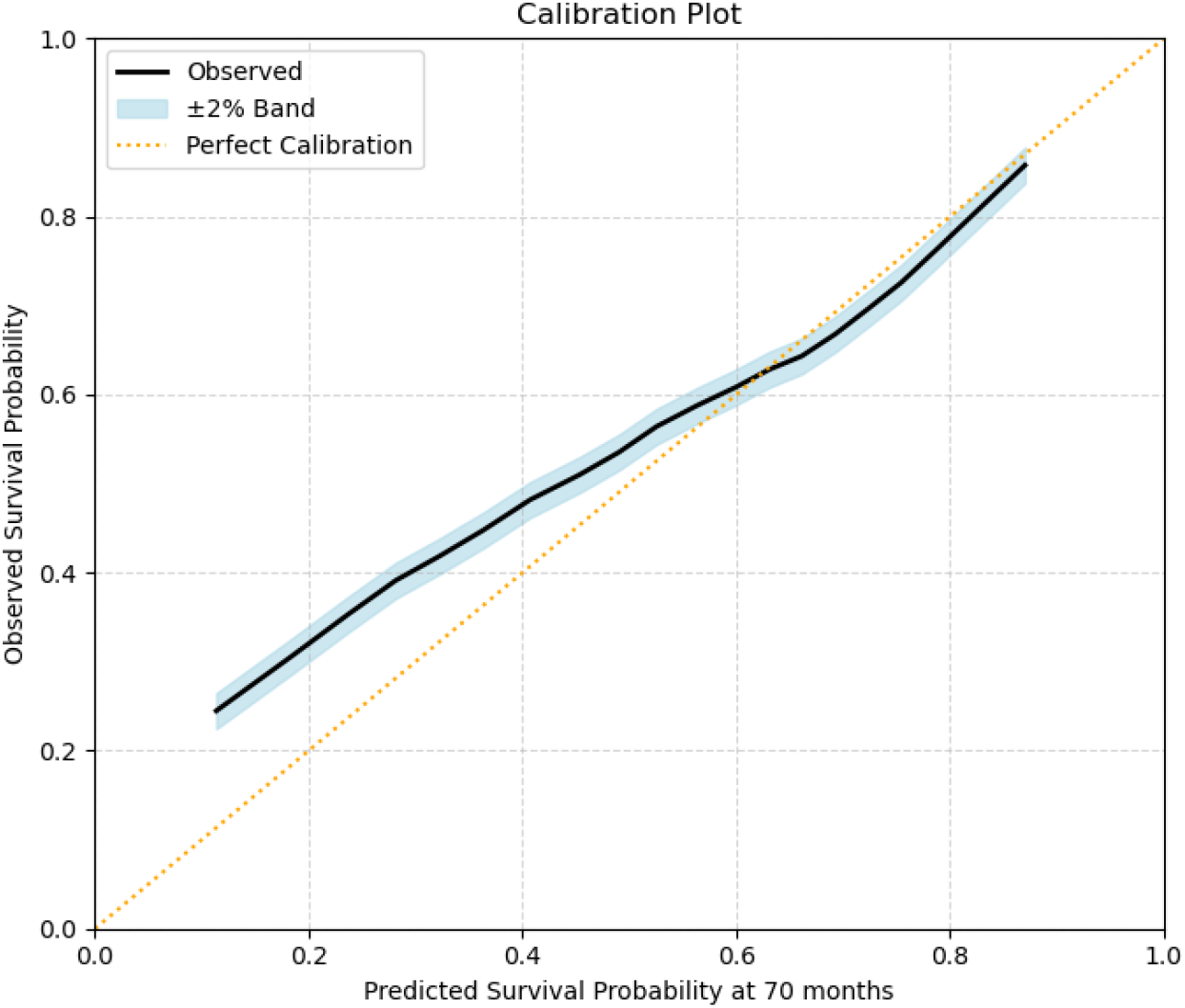
Calibration of the Cox Model in the External Validation Cohort. Observed versus predicted survival probabilities at 70 months for 7,749 patients in the SEER cohort, grouped by deciles of predicted risk. The solid black line represents observed outcomes with a ±2% band (blue shading). The dotted orange line indicates perfect calibration.

### External Validation using SEER Dataset

For external validation, the Cox model was applied to 7,749 patients from the SEER cohort after retraining on overlapping variables. Missing data was infrequent (<5 % of data) and imputed using iterative imputation. The C-index was 0.688. Calibration at 70 months indicated underestimation of survival in higher-risk patients. Because laboratory-based biomarkers (albumin, NLR, platelet count) and comorbidity indices were not available in SEER, the external validation model included only demographic variables (age, sex), tumor characteristics (T, N, M stage, anatomical subsite), and treatment modalities (surgery, radiation, chemotherapy).

Missing data in the SEER cohort was minimal for the variables included in the reduced model. Missing values were handled using iterative imputation.

The reduced Cox model achieved a C-index of 0.688 in the SEER cohort, demonstrating acceptable discrimination despite the absence of biomarker and comorbidity data. Calibration was assessed by grouping patients into deciles of predicted 70-month mortality risk and comparing each group’s predicted survival probability with observed Kaplan–Meier survival. The plot evaluates how closely model-based predictions match real outcomes. The reduced Cox model showed good calibration overall, with most points close to the identity line, although survival was underestimated in the highest-risk deciles.

## Discussion

Using routinely collected EHR data, our validated Cox model demonstrated good performance in predicting survival among patients with advanced HNSCC. The model achieved a C-index of .735 in the internal test set and .688 in partial external validation with SEER. By combining demographics, staging, treatment, comorbidities, and host biomarkers, we developed a tool that can support individualized prognostic estimates, guide treatment discussions, and stratify patients by risk at diagnosis.

Recent systematic reviews of prognostic models in oropharyngeal cancer suggested future models need to be methodologically robust, with large sample size, should attempt to account for missing variable data, and report calibration; they also pointed out the need to explore the prognostic effect of molecular biomarkers [(Dretzke et al., 2024), (Abou-Foul et al., 2024)]. Our analysis directly responded to these suggestions, highlighting several strengths of our analysis.

First, we developed the model in a large, multi-institutional cohort (∼2,200 patients) and externally validated it in SEER, a national registry with far broader coverage. Next, we accounted for missing variable data, such as NLR and serum albumin, by imputation and including a missingness indicator variable in the model. We also reported the calibration curve to ensure transparency. Third, we incorporated SHAP-based interpretability, providing patient-level explanations that extend transparency beyond traditional regression coefficients or survival curves. Finally, we combined clinical variables and inflammatory biomarkers in the model.

Several predictors in our model are consistent with established literature. A high neutrophil-to-lymphocyte ratio was again associated with worse outcomes [(Takenaka et al., 2022), (Kao et al., 2022), (Thayalan et al., 2024)], underscoring its value as a negative prognostic marker. Albumin demonstrated high importance in SHAP analysis despite not being significant in the Cox model, suggesting nonlinear effects are more easily captured by machine learning methods. Higher BMI appeared protective, consistent with evidence that nutritional reserve supports treatment tolerance [(Wu et al., 2022), (Hobday et al., 2023), (Hicks et al., 2018), (Ma et al., 2023)]. Among treatment factors, surgery was the strongest positive predictor of survival, aligning with the clinical observation that surgically resectable disease has a better prognosis, while the impact of chemotherapy and radiation is less consistent. These findings suggest that non-linear models may capture prognostic relationships beyond those identified by traditional regression.

This framework has clear clinical implications. The model provides patient-specific survival estimates based on data from thousands of cases, enabling patients and clinicians to understand how prognosis varies by treatment choice. It also refines risk stratification: patients with favorable stage disease but poor biomarker profiles can be reclassified into higher-risk groups, highlighting the importance of host factors alongside tumor stage [Chen et al., 2022]. Our clinical prognosis modeling can facilitate discussions with patients on prognosis and expectations for treatment, particularly in situations where treatment decisions are difficult to make [Zhang et al., 2023]. Clinicians can utilize our model or a similar model trained with institutional data as a clinical decision support tool, rather than relying on published data that may not be reflective of the patient population of the practice setting.

A Cox model developed by Hoesseini et al. incorporated performance status, socioeconomic factors, and HPV/p16 status and achieved C-indices of approximately 0.67 to 0.71 [Hoesseini et al., 2021]. The multimodal model by Mansouri et al. combined clinical variables with CT radiomics and dosiomics features and reported C-indices up to 0.73 [Mansouri et al., 2024]; despite using only routinely available clinical variables, comorbidities, and laboratory biomarkers, our model achieved similar discrimination (C-index 0.735), underscoring its practicality for real-world implementation.

Because the approach is built on the OMOP common data model, it could be integrated into institutional EHR systems for automated risk stratification, embedded in clinical workflows, and tailored to site-specific populations [Park et al., 2023]. In practice, such a tool could identify patients who may benefit from closer surveillance, early nutritional or palliative referral, or more intensive supportive care, while reserving resources for lower-risk groups. Cutoffs for risk categories could also be adapted to institutional resource availability, trading sensitivity for specificity depending on whether the priority is to minimize missed high-risk patients or to limit false alarms.

This study has limitations. HPV status was not available, which is a major limitation for oropharyngeal cancer specifically [(Fakhry et al., 2008), (Khanna et al., 2020)]. Our model was developed for advanced HNSCC across multiple subsites, where HPV plays a less consistent role [(Li et al., 2018), (BOSCOLO-RIZZO et al., 2013)], but standardized HPV capture across registries and EHRs remains a major barrier for future model development. In addition, the neutrophil-to-lymphocyte ratio had substantial missingness, which we addressed through imputation and sensitivity analyses, but with potential residual bias. External validation in SEER was restricted to demographics, staging, subsite, and treatment variables, excluding biomarkers and comorbidities, though the model still achieved a C-index of 0.68 under these constraints. Taken together, these limitations highlight the need for more complete data capture in real-world datasets, particularly HPV status and biomarkers, and for further validation in diverse health systems.

## Conclusion

In this prognostic modeling study of more than 2,000 patients with advanced head and neck squamous cell carcinoma, a machine learning model using routinely collected clinical and biomarker data achieved good discrimination and calibration for survival prediction, with partial validation in an external registry. These findings suggest that machine learning can integrate diverse clinical features to provide individualized prognostic estimates, support treatment discussions, and refine risk stratification in head and neck cancer. Further validation across diverse populations and inclusion of molecular data are needed before clinical implementation.

## Data Availability

All data produced in the present study are available upon reasonable request to the authors

